# Evaluation of the performance of SARS-CoV-2 antibody assays for the longitudinal population-based study of COVID-19 spread in St. Petersburg, Russia

**DOI:** 10.1101/2021.04.05.21254712

**Authors:** Anton Barchuk, Daniil Shirokov, Mariia Sergeeva, Rustam Tursun-zade, Olga Dudkina, Varvara Tychkova, Lubov Barabanova, Dmitriy Skougarevskiy, Daria Danilenko

**Affiliations:** European University at St. Petersburg, Shpalernaya Ulitsa, 1, St. Petersburg, Russia, 191187; Clinic “Scandinavia” (LLC Ava-Peter), Ilyushina Ulitsa, 4-1, St. Petersburg, Russia, 197372; Smorodintsev Research Institute of Influenza, Ulitsa Professora Popova, 15/17, St. Petersburg, Russia, 197022; Petrov National Research Medical Center of Oncology, Pesochny, Leningradskaya Ulitsa, 68, St. Petersburg, Russia, 197758; ITMO University, Kronverksky Prospekt, 49, St. Petersburg, Russia, 197101

**Keywords:** SARS-CoV-2, COVID-19, seroepidemiologic study, SARS-CoV-2 infection antibody testing

## Abstract

**Background:** An evident geographical variation in the SARS-CoV-2 spread requires seroprevalence studies based on local tests with robust validation against already available antibody tests and neutralization assays. This report summarizes the evaluation of antibody tests used in the representative population-based serological study of SARS-CoV-2 in Saint Petersburg, Russia.

**Methods:** We used three different antibody tests throughout the study: chemiluminescent microparticle immunoassay (CMIA) Abbott Architect SARS-CoV-2 IgG, Enzyme-linked immunosorbent assay (ELISA) CoronaPass total antibodies test, and ELISA SARS-CoV-2-IgG-EIA-BEST. Clinical sensitivity was estimated with the SARS-CoV-2 PCR test as the gold standard and specificity in pre-pandemic sera samples using the cut-off recommended by manufacturers. Paired and unpaired serum sets were used. Measures of concordance were also calculated in the seroprevalence study sample against the microneutralization test (MNA).

**Findings:** Sensitivity was equal to 91.1% (95% CI: 78.8–97.5) and 90% (95% CI: 76.4–96.4) for ELISA Coronapass and ELISA Vector-Best respectively. It was equal to 63.1% (95% CI (50.2–74.7) for CMIA Abbott. Specificity was equal to 100% for all the tests. Comparison of ROCs for three tests has shown lower AUC for CMIA Abbott, but not for ELISA Coronapass and CMIA Abbott. The cutoff SC/O ratio of 0.28 for CMIA-Abbott resulted in a sensitivity of 80% at the same full level of specificity. In less than one-third of the population-based study participants with positive antibody test results, we detected neutralizing antibodies in titers 1:80 and above. There was a moderate correlation between antibody assays results and MNA.

**Interpretation:** Our validation study encourages the use of local antibody tests for population-based SARS-CoV-2 surveillance and sets the reference for the seroprevalence correction. Available tests are sensitive enough to detect antibodies in most individuals with previous positive PCR tests with a follow-up of more than 5 months. The Abbott Architect SARS-CoV-2 IgG’s sensitivity can be significantly improved by incorporating a new cut-off. Relying on manufacturers’ test characteristics for correction of reported prevalence estimates may introduce bias to the study results.

**Funding:** Polymetal International plc

## Introduction

SARS-CoV-2 seroprevalence studies have proved to be a valuable tool in the assessment of the COVID-19 pandemic dynamics [1–3]. However, researchers have raised several concerns regarding the non-response bias, the non-representative sampling, and the use of non-validated tests for SARS-CoV-2 antibodies detection [4]. The first two issues can be addressed by appropriate study design and population sampling strategies [5; 6]. The latter problem requires the assessment of sensitivity and specificity of antibody assays to guide the correction of serological study results [7–11]. Additional requirements for antibody tests are being lodged for longitudinal serologic studies, but manufacturers rarely report the evaluation of the long-term test performance, e.g., decreased long-term sensitivity is an issue that could bias the evaluation of immune response durability [12; 13]. An evident geographical variation in the spread of SARS-CoV-2 requires local seroprevalence studies [14]. The use of locally available tests from national manufacturers can be convenient but often comes at the expense of test performance [3]. Local tests require robust validation against the benchmark of already available antibody tests and microneutralization assays using sera samples from cases confirmed by PCR and from healthy donors.

Long-term performance of antibody tests is key to understanding the antibody kinetics. Previous validation studies have shown significant changes in the sensitivity of certain antibody tests in the longer follow-up[15]. This report summarizes the validation of antibody tests used in the representative population-based serological study of SARS-CoV-2 in St. Petersburg, Russia, a densely populated city with more than 5 million inhabitants making it the fourth largest city in Europe [6]. We aimed at establishing test specificity (Sp) and sensitivity (Se) to correct estimates obtained through population-based seroprevalence study.

## Materials and methods

Information about the corresponding seroprevalence study in St. Petersburg is available in the detailed report [6]. In brief, we obtained a representative population sample through phone by using random-digit dialling. The study started in May 2020. The first report was publicly available in August 2020. This study is underway and includes three rounds of blood sample collection with more than 2,500 participants.

### Antibody tests

We used three different antibody tests throughout the study: 1) chemiluminescent microparticle immunoassay Abbott Architect SARS-CoV-2 IgG on the Abbott ARCHITECT® i2000sr platform (Abbott Laboratories, Chicago, USA), detecting immunoglobulin class G (IgG) antibodies to the nucleocapsid protein of SARS-CoV-2 with the signal/cut-off (S/CO) ratio of 1.4 for positivity (CMIA Abbot; www.fda.gov/media/137383/download).; 2) enzyme-linked immunosorbent assay CoronaPass total antibodies test (Genetico, Moscow, Russia) based on recombinant receptor binding domain of the spike protein of SARS-CoV-2 (Department of Microbiology, Icahn School of Medicine at Mount Sinai, New York, NY, USA), detecting total antibodies with the S/CO ratio of 1.0 for positivity (ELISA Coronapass, pass.genetico.ru); 3) enzyme-linked immunosorbent assay SARS-CoV-2-IgG-EIA-BEST by Vector-Best, Novosibirsk, Russia also detecting IgG antibodies to the spike protein of SARS-CoV-2 with the S/CO ratio of 1.1 for positivity.(ELISA Vector-Best, vector-best.ru/en/prod/index.php?SECTION_ID=2724).

Test performance for CMIA Abbott is available from the manufacturer materials (Se=100%, Sp=99.6%). It was also evaluated in numerous independent studies [9; 16–18]. ELISA Coronapass test manufacturer provides information on its official website (Se=98.7%, Sp=100%). ELISA Vector-Best sensitivity and specificity is reported reported in one study published in Russian (Se=100%, Sp=99.8%) [19].

We used CMIA Abbott and ELISA Coronapass for the first publication of a population-based seroprevalence study in May-June 2020 [6]. The first round of the study analysis showed that the use of ELISA Coronapass results in a slightly higher seroprevalence estimation. It was the initial reason for the independent validation of the test performance.

In the sensitivity assessment, we also used CMIA Elecsys Anti-SARS-CoV-2, manufactured by Roche Diagnostics GmbH, Mannheim, Germany (CMIA Roche, S/CO ratio of 1.0 for positivity, Se=99.5%, Sp=99.8%; diagnostics.roche.com/ru/ru/products/params/elecsys-anti-sars-cov-2.html).

### Samples for the sensitivity and specificity assessment

For the sensitivity assessment, we obtained 92 serum samples collected from the staff of “Scandinavia” clinic as part of routine antibody monitoring carried out in the clinic. Serum samples were obtained for individuals who reported positive PCR tests for SARS-CoV-2 RNA. The median time between a positive PCR test result and blood draw for antibody test was equal to 21 weeks [IQR: 19.9-22.5].

For the specificity assessment, pre-pandemic human sera samples were obtained from healthy blood donors collected by Smorodintsev Research Institute of Influenza through routine influenza surveillance in different cities across Russia. Sera samples are collected twice a year to assess the herd immunity against influenza A and B viruses. Written informed consent was obtained in all cases of blood donation and further sample preparation. We used 93 remnant sera samples from herd immunity studies that were kept frozen at −80°C.

### Neutralization test

Neutralization test was performed for 365 samples that were positive for binding antibody samples and for 74 negative samples. TCID_50_ based microneutralization test (MNA) was used to detect and titrate neutralizing antibodies [20]. First, serum samples were heated for 30 min at 56°C to avoid complement-linked reduction of the viral activity. This was followed by the preparation of serial two-fold dilutions starting from 1:10 in 60 uL volume (tested in triplicate) of each serum specimen in culture medium (Alpha MEM containing antibiotics and 2% heat-inactivated fetal bovine serum). Each dilution was mixed at equal volume with the live SARS-CoV-2 virus (60 uL, containing 25 TCID_50_/50uL) and incubated for 60 minutes at 37°C in plastic microplates. Then 100 μl of the mix was transferred into 96-well microplates with monolayer Vero cells. The plates were incubated at 37°C in a 5% CO2 atmosphere. Readings were evaluated 5 to 6 days later and neutralization was recorded if 100% of the cells in the well were preserved (no visible plaques or cytopathic effect). Serum neutralizing titer was expressed as the inverse of the higher serum dilution that exhibited neutralizing activity. All experiments were performed in a BSL3 laboratory.

### Statistical analyses

Clinical sensitivity was estimated with the SARS-CoV-2 PCR test as the gold standard and specificity in pre-pandemic sera samples. We used 46 PCR positive samples and 41 pre-pandemic serum samples to cross-validate all three tests (cross-validation sample). In the independent test validation (full-validation sample), different sets of non-paired samples (60 PCR positive samples and 65 pre-pandemic samples for CMIA Abbott, 48 and 93 for ELISA Vector-Best, 60 and 92 for ELISA-Coronapass) were used. We report the validation study results based on the manufacturers’ recommended S/CO ratios, except for CMIA-Abbott, we use both the S/CO ratio of 1.4 and a more sensitive S/CO ratio of 1.0. Compared with the neutralization test results, the percent agreement (positive, negative and overall), the concordance (Cohen’s Kappa coefficient along with the prevalence index (P-index), bias index (B-index), and prevalence and bias-adjusted kappa — PABAK), and Spearman correlation coefficients were calculated [21].

The estimates are given with the 95% confidence intervals (CI) where appropriate. `R` statistical software (v.4.0.2; `R` Foundation for Statistical Computing, Vienna, Austria) was used for data analysis and presentation. Exact binomial confidence limits were calculated for test sensitivity, specificity, and percent agreement (negative — NPA, positive — PPA, and overall — OPA) applying the `epi.tests` function from the `epiR package`. ROC curves were constructed, and the corresponding area under the curve (AUC) was calculated for three tests using the pROC package. CIs for AUCs were calculated with 2000 bootstrap replicates. The bootstrap test was used to compare ROC curves: a test for paired curves was used in the cross-validation sample and non-paired in the full-validation sample. Confidence intervals were calculated for Spearman’s correlation with the `spearmanCI` package.

To report CIs for concordance measures — bias, prevalence, and kappa — we used the bootstrap percentile method (the `epi.kappa` function from the `epiR` package). Formal criteria (eg., Landis and Koch) for Cohen’s Kappa interpretation were not used.

### Data sharing

Study data and code is available online (https://github.com/eusporg/spb_covid_study20/tree/master/validation_of_covid_tests).

### Ethical considerations and study registration

The study was approved by the Research Planning Board of European University at St. Petersburg (on May 20, 2020) and the Ethic Committee of the Clinic “Scandinavia” (on May 26, 2020). All research was performed in accordance with the relevant guidelines and regulations. Informed consent was obtained from all participants of the study. The study was registered with the following identifiers: Clinicaltrials.gov (NCT04406038, submitted on May 26, 2020, date of registration — May 28,2020) and ISRCTN registry (ISRCTN11060415, submitted on May 26, 2020, date of registration — May 28,2020). The amendment, which included a test validation study, was approved by the Ethics Committee of the Clinic “Scandinavia” on October 2, 2020. We used blood samples collected from the clinic “Scandinavia” staff as a part of the routine antibody monitoring for which the hospital obtained a separate informed consent. Additional oral consent was obtained to use collected samples for the validation study. Personal information was not linked to serum samples in the validation study except for the date of positive PCR for SARS-CoV-2.

## Results

Based on cross-validation results, the sensitivity was equal to 91.1% (95% CI: 78.8–97.5) for ELISA Coronapass and to 89.1% (95% CI: 76.4–96.4) for ELISA Vector-Best respectively. It was not significantly different from the CMIA Roche — 89.1% (95% CI: 76.4–96.4). However, the sensitivity of CMIA Abbott was equal to 63.1% (95% CI 50.2–74.7). It was slightly higher with the cutoff SC/O ratio of 1.0 — 70.7% (95% CI: 57.3–81.9). Specificity was equal to 100% for all the tests (see Table 1).

**Table 1.** Sensitivity and specificity of tests based on cross-validatation (paired serum samples)

| Test | N | Sensitivity, % (95% CI) | N | Specificity, % (95% CI) |
| --- | --- | --- | --- | --- |
| CMIA Abbott 1.0 | 46 | 70.7 (57.3-81.9) | 41 | 100 (88.1-100) |
| CMIA Abbott 1.4 | 46 | 63.1 (50.2-74.7) | 41 | 100 (84.6-100) |
| CMIA Roche | 46 | 89.1 (76.4-96.4) | — | — |
| ELISA Coronapass | 46 | 91.1 (78.8-97.5) | 41 | 100 (91.6-100) |
| ELISA Vector-Best | 46 | 89.1 (76.4-96.4) | 41 | 100 (91.4-100) |

Using non-paired samples in the validation of the tests did not dramatically change the sensitivity and point estimates but narrowed confidence intervals (see Supplementary Table S1).

Although the comparison of ROCs for three tests has shown that AUC is lower for CMIA-Abbott, the difference was not dramatic — 0.96 (95% CI: 0.90–1.00) for ELISA Coronapass and 0.90 (95% CI: 0.82–0.97) for CMIA Abbott, the difference was only significant when paired samples were compared (see Table 2 and Table S2).

**Table 2.**
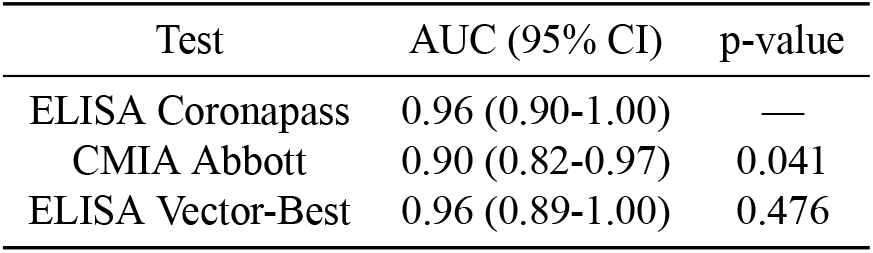
Area under the ROC (AUC) of tests based on cross-validation (paired serum samples)

| Test | AUC (95% CI) | p-value |
| --- | --- | --- |
| ELISA Coronapass | 0.96 (0.90-1.00) | — |
| CMIA Abbott | 0.90 (0.82-0.97) | 0.041 |
| ELISA Vector-Best | 0.96 (0.89-1.00) | 0.476 |

The cutoff SC/O ratio of 0.28 for CMIA-Abbott resulted in a sensitivity of 80% at the same full level of specificity. The other test thresholds were optimal based on the ROC analysis of cross-validation and non-paired samples (see Figure 1).

**Figure 1.**
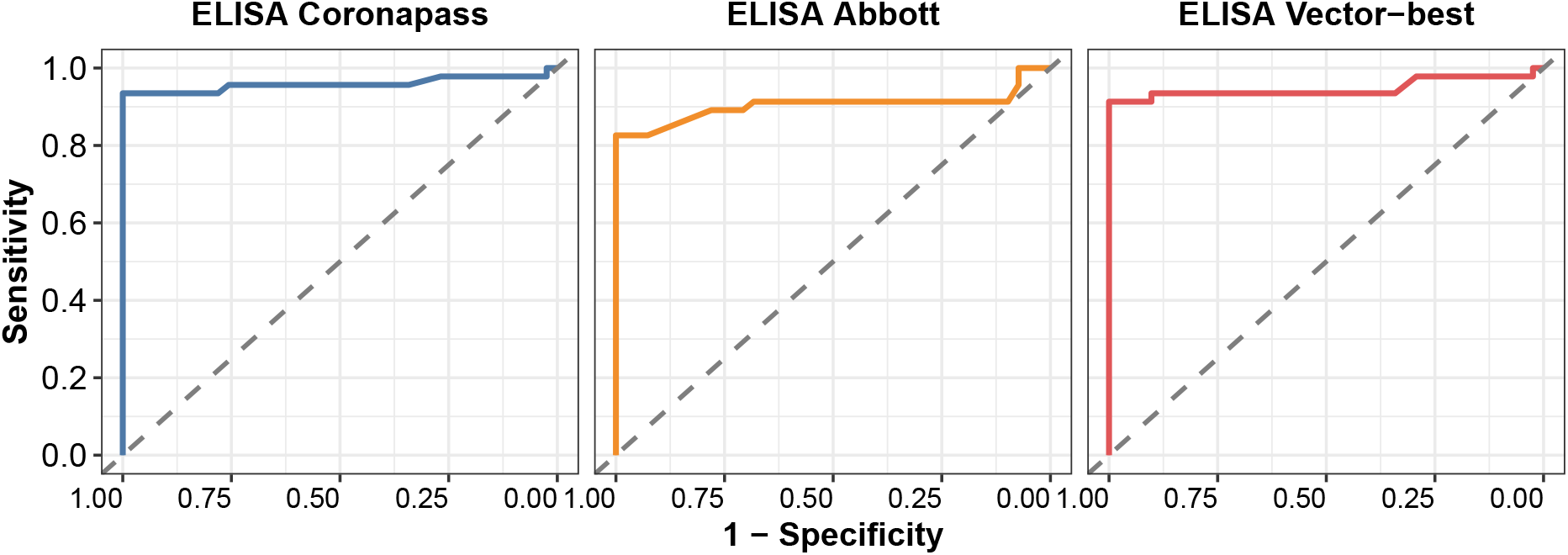
ROCs for three test against PCR test results

In less than one-third of the population-based study participants with positive antibody total IgG test results, we detected neutralizing antibodies in titers 1:80 and above. NPA was between 28.0% (95% CI: 22.0–34.5) for CMIA Abbott (cut-off 1.4) and 20.8% (95% CI: 16.6–25.5) for ELISA Vector-best. NPA between MNA with a cut-off at titers 1:20 was between 74.4% (95% CI: 68.0–80.2) for CMIA Abbott (cut-off 1.4) and 64.2% (95% CI: 58.9–69.3) for ELISA Vector-Best (see Table 3).

**Table 3.** Percent agreement between antibody tests and neutralizing antibody tests using two cut-offs (titers 1:20 and 1:80)

| Test | MNA100 cut-off | PPA (95% CI) | NPA (95% CI) | OPA (95% CI) |
| --- | --- | --- | --- | --- |
| CMIA Abbott 1.0 | 20 | 76.5 (70.0-82.2) | 72.0 (65.8-77.6) | 74.0 (69.7-78.1) |
| CMIA Abbott 1.4 | 20 | 72.8 (66.5-78.5) | 74.4 (68.0-80.2) | 73.6 (69.2-77.6) |
| ELISA Coronapass | 20 | 91.1 (84.6-95.5) | 65.8 (60.3-71.0) | 72.9 (68.5-77.0) |
| ELISA Vector-Best | 20 | 100 (96.3-100) | 64.2 (58.9-69.3) | 72.2 (67.8-76.4) |
| CMIA Abbott 1.0 | 80 | 95.5 (91.6-97.9) | 25.9 (20.5-32.0) | 57.6 (52.9-62.3) |
| CMIA Abbott 1.4 | 80 | 94.7 (91.0-97.3) | 28.0 (22.0-34.5) | 62.6 (57.9-67.2) |
| ELISA Coronapass | 80 | 96.7 (91.9-99.1) | 21.2 (16.8-26.1) | 42.4 (37.7-47.1) |
| ELISA Vector-Best | 80 | 100 (96.3-100) | 20.8 (16.6-25.5) | 38.5 (33.9-43.2) |

The measures of concordance are presented in Table S3. There was a moderate correlation between antibody assays results and MNA: 0.65 (95% CI: 0.59–0.71) for CMIA Abbott, 0.60 (95% CI: 0.54–0.67) for ELISA Coronapass, and 0.76 (95% CI: 0.72–0.81) for ELISA Vector-Best (see Figure 2).

**Figure 2.**
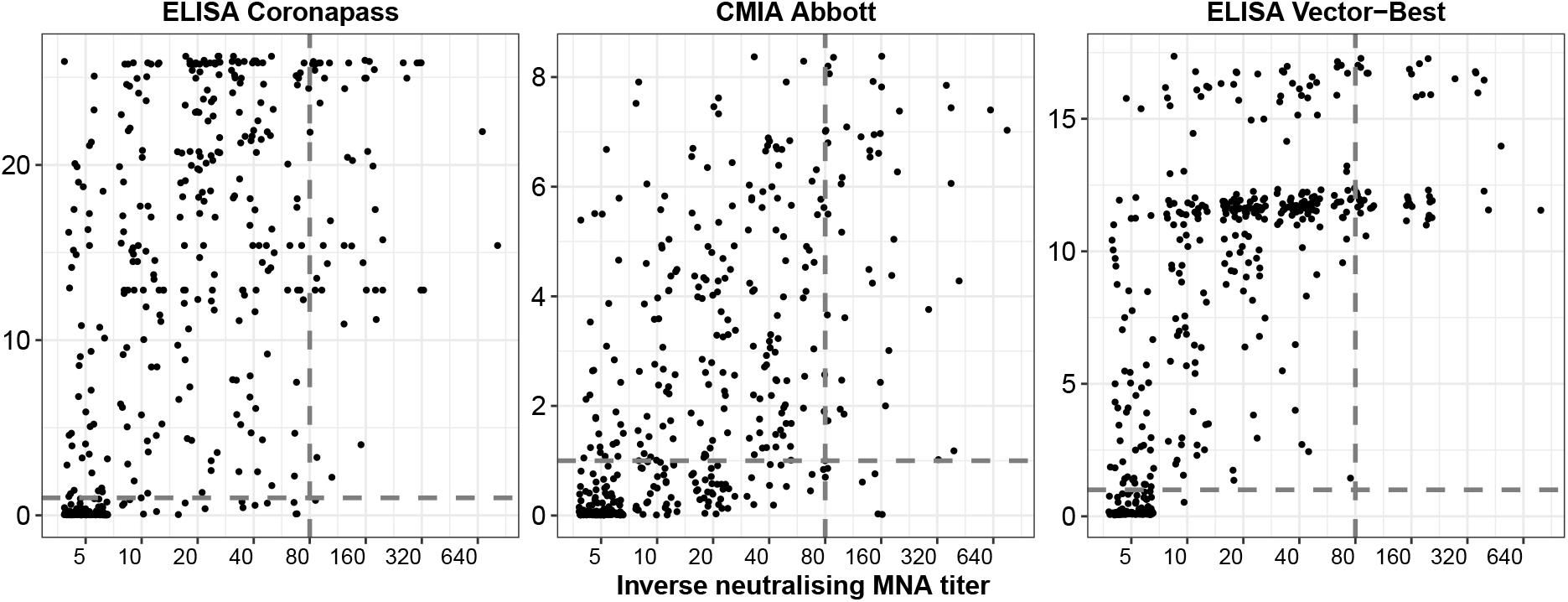
Correlation between antibody tests results and neutralization test

## Discussion

Our validation study encourages the use of local antibody tests for population-based SARS-CoV-2 surveillance and sets the reference for the seroprevalence correction. However, it discourages the use of Abbott Architect SARS-CoV-2 IgG for population-based seroprevalence SARS-CoV-2 surveillance. The decrease in test sensitivity (63% with a cut-off of 1.4) is likely to be related to the long period between infection and blood sampling compared to the manufacturer’s validation studies (it was on average 21 weeks after the reported positive PCR test). Previous studies also found similar results for samples taken at a longer follow-up [13; 15]. These results may also explain the decline in antibody presence in longitudinal studies that may be incorrectly interpreted by the short-term immune response to SARS-CoV-2 [22].

Based on our validation study, the Abbott Architect SARS-CoV-2 IgG’s sensitivity can be significantly improved by incorporating a new cut-off. Moving the S/CO ratio from 1.4 or 1.0 for positivity to 0.28 improved sensitivity from 63% or 71% to 80%, respectively, without loss in specificity.

Locally available ELISA tests and Roche assays are sensitive enough to detect antibodies in most individuals with previous positive PCR tests. Tests were negative for approximately 10% of participants with positive PCR, with any antibody tests. From the available data, it is impossible to conclude whether test results are false-negative antibody, false-positive PCR, or that infection did not result in the antibody response. Further studies are also needed to look into other mechanisms of the immune response to SARS-CoV-2 [23].

The results for the assessment of concordance between binding and neutralizing antibodies tests should be interpreted with caution. Less than a third of samples positive for binding antibodies were also positive in the virus neutralization test (with 1:80 titer as a threshold). However, this does not mean that they are susceptible to SARS-CoV-2 virus re-infections. Case reports about second episode of COVID-19 infection in the presence of neutralizing antibodies were also published [24]. The follow-up time plays an essential role in neutralization test results, but negative tests do not rule out other protection mechanisms. Recent studies showed that a positive antibody test is a powerful predictor of lowered risk of SARS-CoV-2 reinfection measures as a positive PCR test [25; 26].

This finding has an important practical implication. Relying on test characteristics provided by manufacturers for correction of reported prevalence estimates introduces additional bias to the study. If the test reported prevalence is 5%, then with the test sensitivity of 65% and specificity of 100%, the true prevalence would be equal to approximately 8% [27]. If the crude prevalence based on the same test is 40%, then the true prevalence would be around 62%. The magnitude of bias in the population with a significant proportion of individuals with antibodies would limit the ability to detect herd immunity threshold.

This study has several limitations. PCR test results that were chosen as a golden standard, were carried out in different laboratories. Although official test certificates that are registered in the national database were provided, false-positive results cannot be ruled out [28]. But as mentioned above, PCR false-positivity is likely to underestimate antibody test sensitivity in this study. Neutralization test results should be considered with caution as well [29]. Such tests can be considered as a surrogate marker of protection from reinfection, but this association needs to be explored in population-based epidemiological studies [15].

In conclusion, this validation study provides a reference that can be used in further seroprevalence reports to correct the results based on test sensitivity and specificity. Choice of the test for longitudinal surveillance is critical for making conclusions about the spread of SARS-CoV-2 and the durability of the immune response. Local tests should be rigorously evaluated for seroprevalence studies because the benefits of using properly validated tests are not only financial or related to matters of convenience. They may provide more accurate and unbiased assessments for the course of the pandemic.

## Data Availability

Study data and code is available online.

https://github.com/eusporg/spb_covid_study20/tree/master/validation_of_covid_tests

## Authors’ contributions

AB, DS, DD and DSk conceived the study. AB drafted the first version of the manuscript. DD and VT performed Vector-Best ELISA assays, MS performed the neutralization assays. AB, DSk, RT and OD did data analyses. All authors participated in the study design, contributed to the interpretation of data and to drafting sections of the manuscript. All authors read and approved the final manuscript.

## Acknowledgements

We acknowledge personal support from Vitaly Nesis (Chief Executive Officer, Polymetal International, plc). We thank Alla Samoletova (European University at St. Petersburg) for administrative support and management of the study. The authors also thank Alyona Zheltukhina from Smorodintsev Research Institute of Influenza for excellent technical assistance. We thank the nurses, general practitioners, laboratory and administrative personnel of the Clinic “Scandinavia” and Smorodintsev Research Institute of Influenza.

## Supplementary materials

**Table S1.** Sensitivity and specificity of tests based on full validation sample (non-paired serum samples)

| Test | N | Sensitivity, % (95% CI) | N | Specificity, % (95% CI) |
| --- | --- | --- | --- | --- |
| CMIA Abbott 1.0 | 65 | 74.7 (64.3-83.4) | 60 | 100 (90.7-100) |
| CMIA Abbott 1.4 | 65 | 67.7 (57.4-76.9) | 60 | 100 (88.1-100) |
| CMIA Roche | 61 | 86.9 (75.8-94.2) | - | - |
| ELISA Coronapass | 92 | 92.0 (84.8-96.5) | 60 | 100 (93.2-100) |
| ELISA Vector-Best | 93 | 93.9 (87.3-97.7) | 48 | 100 (91.6-100) |

**Table S2.**
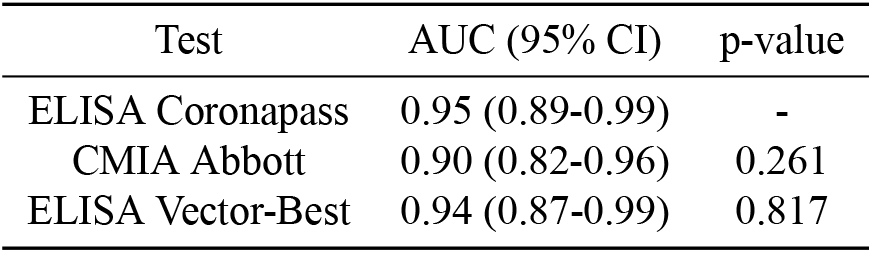
Area under the ROC (AUC) of tests based on full validation sample (non-paired serum samples)

**Table S3.**
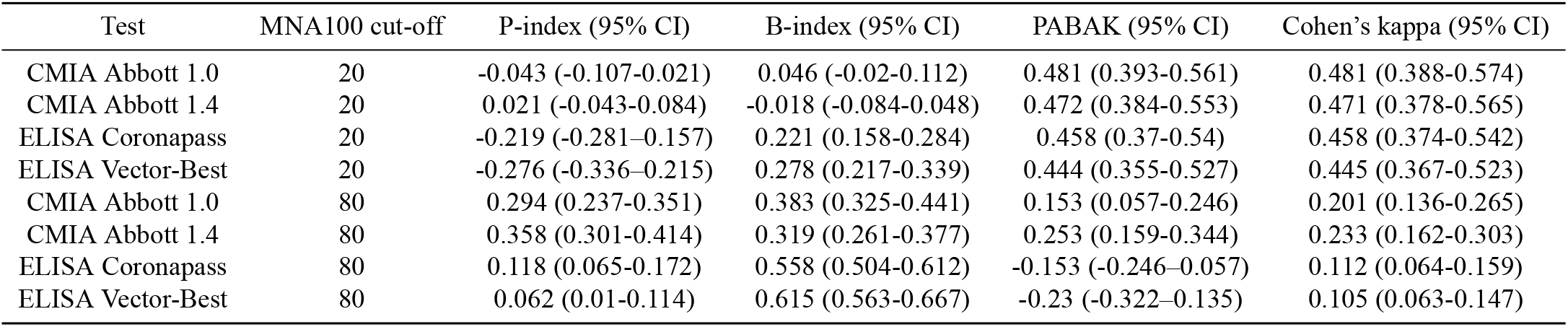
Concordance measures between antibody tests and neutralizing antibody tests using two cut-offs (titers 1:20 and 1:80)

## References

1. Goudsmit J. The paramount importance of serological surveys of SARS-CoV-2 infection and immunity. European Journal of Epidemiology. 2020:1.

2. Gudbjartsson DF, Norddahl GL, Melsted P, Gunnarsdottir K, Holm H, Eythorsson E, et al. Humoral immune response to SARS-CoV-2 in Iceland. New England Journal of Medicine. 2020;383(18):1724–1734.

3. Pollán M, PérezGómez B, Pastor-Barriuso R, Oteo J, Hernán MA, Pérez-Olmeda M, et al. Prevalence of SARS-CoV-2 in Spain (ENE-COVID): a nationwide, population-based seroepi demiological study. The Lancet. 2020;396(10250):535–544.

4. Bennett ST, Steyvers M. Estimating COVID-19 antibody seroprevalence in Santa Clara County, California. A re-analysis of Bendavid et al. MedRxiv. 2020.

5. Mosha N, Aluko O, Todd J, Machekano R, Young T. Analytical methods used in estimating the prevalence of HIV/AIDS from demographic and cross-sectional surveys with missing data: a systematic review. BMC Medical Research Methodology. 2020;20(1):1–10.

6. Barchuk A, Skougarevskiy D, Titaev K, Shirokov D, Raskina Y, Novkunkskaya A, et al. Seroprevalence of SARS-CoV-2 antibodies in Saint Petersburg, Russia: a population-based study. MedRxiv. 2020.

7. Harritshøj LH, Gybel-Brask M, Afzal S, Kamstrup PR, Jørgensen CS, Thomsen MK, et al. Comparison of sixteen serological SARS-CoV-2 immunoassays in sixteen clinical laboratories. Journal of Clinical Microbiology. 2021.

8. Eyre DW, Lumley SF, O’Donnell D, Stoesser NE, Matthews PC, Howarth A, et al. Stringent thresholds in SARS-CoV-2 IgG assays lead to under-detection of mild infections. BMC infectious diseases. 2021;21(1):1–10.

9. Gutiérrez-Cobos A, de Frutos SG, García DD, Lara EN, Carrión AY, García-Rodrigo LF, et al. Evaluation of diagnostic accuracy of 10 serological assays for detection of SARS-CoV-2 antibodies. European Journal of Clinical Microbiology & Infectious Diseases. 2020:1–7.

10. Weidner L, Gänsdorfer S, Unterweger S, Weseslindtner L, Drexler C, Farcet M, et al. Quantification of SARS-CoV-2 antibodies with eight commercially available immunoassays. Journal of Clinical Virology. 2020;129:104540.

11. Wehrhahn MC, Brown S, Newcombe JP, Chong S, Evans J, Figtree M, et al. An evaluation of 4 commercial assays for the detection of SARS-CoV-2 antibodies in a predominantly mildly symptomatic low prevalence Australian population. Journal of Clinical Virology. 2021:104797.

12. Dittadi R, Afshar H, Carraro P. Two SARS-CoV-2 IgG immunoassays comparison and time-course profile of antibodies response. Diagnosguttic Microbiology and Infectious Disease. 2021;99(4):115297.

13. Bal A, Trabaud MA, Fassier JB, Rabilloud M, Saker K, Langlois-Jacques C, et al. Six-month antibody response to SARS-CoV-2 in healthcare workers assessed by virus neutralization and commercial assays. Clinical Microbiology and Infection. 2021.

14. Rostami A, Sepidarkish M, Leeflang M, Riahi SM, Shiadeh MN, Esfandyari S, et al. SARS-CoV-2 seroprevalence worldwide: a systematic review and meta-analysis. Clinical Microbiology and Infection. 2020.

15. Muecksch F, Wise H, Batchelor B, Squires M, Semple E, Richardson C, et al. Longitudinal Serological Analysis and Neutralizing Antibody Levels in Coronavirus Disease 2019 Convalescent Patients. The Journal of Infectious Diseases. 2020.

16. Chew KL, Tan SS, Saw S, Pajarillaga A, Zaine S, Khoo C, et al. Clinical evaluation of serological IgG antibody response on the Abbott Architect for established SARS-CoV-2 infection. Clinical Microbiology and Infection. 2020;26(9):1256–e9.

17. Batra R, Olivieri LG, Rubin D, Vallari A, Pearce S, Olivo A, et al. A comparative evaluation between the Abbott Panbio™ COVID-19 IgG/IgM rapid test device and Abbott Architect™ SARS -CoV-2 IgG assay. Journal of Clinical Virology. 2020;132:104645.

18. Harley K, Gunsolus IL. Comparison of the Clinical Performances of the Abbott Alinity IgG, Abbott Architect IgM, and Roche Elecsys Total SARS-CoV-2 Antibody Assays. Journal of Clinical Microbiology. 2020;59(1).

19. Kuvshinova I, Nekrasov B, Livitskaia N, Molodykh S, Rukavishnikov M. Sensitivity and specificity of JSC Vector-Best assays for immunoglobulin of different classes to SARS-CoV-2 [Tchuvstvitel’nost’ i specifichnost’ naborov reagentov AO «VektorBest" dlja vyjavlenija immunoglobulinov raznyx klassov k SARS-CoV-2]. Spravochnik Zaveduyuschego KDL. 2020;(10):27–32.

20. Manenti A, Maggetti M, Casa E, Martinuzzi D, Torelli A, Trombetta CM, et al. Evaluation of SARS-CoV-2 neutralizing antibodies using a CPE-based colorimetric live virus micro-neutralization assay in human serum samples. Journal of medical virology. 2020;92(10):2096–2104.

21. Byrt T, Bishop J, Carlin JB. Bias, prevalence and kappa. Journal of clinical epidemiology. 1993;46(5):423–429.

22. Stefanelli P, Bella A, Fedele G, Fiore S, Pancheri S, Benedetti E, et al. Longevity of seropositivity and neutralizing titers among SARS-CoV-2 infected individuals after 4 months from baseline: a population-based study in the province of Trento. medRxiv. 2020.

23. Wheatley AK, Juno JA, Wang JJ, Selva KJ, Reynaldi A, Tan HX, et al. Evolution of immune responses to SARS-CoV-2 in mild-moderate COVID-19. Nature communications. 2021;12(1):1–11.

24. Zhang J, Ding N, Ren L, Song R, Chen D, Zhao X, et al. COVID-19 reinfection in the presence of neutralizing antibodies. National Science Review. 2021.

25. Lumley SF, O’Donnell D, Stoesser NE, Matthews PC, Howarth A, Hatch SB, et al. Antibody status and incidence of SARS-CoV-2 infection in health care workers. New England Journal of Medicine. 2021;384(6):533–540.

26. Harvey RA, Rassen JA, Kabelac CA, Turenne W, Leonard S, Klesh R, et al. Association of SARS-CoV-2 Seropositive Antibody Test With Risk of Future Infection. JAMA Internal Medicine.2021.

27. Reiczigel J, Földi J, ózsvári L. Exact confidence limits for prevalence of a disease with an imperfect diagnostic test. Epidemiology & Infection. 2010;138(11):1674–1678.

28. Skittrall JP, Wilson M, Smielewska AA, Parmar S, Fortune MD, Sparkes D, et al. Specificity and positive predictive value of SARS-CoV-2 nucleic acid amplification testing in a low-prevalence setting. Clinical Microbiology and Infection. 2020.

29. Nie J, Li Q, Wu J, Zhao C, Hao H, Liu H, et al. Establishment and validation of a pseudovirus neutralization assay for SARS-CoV-2. Emerging microbes & infections. 2020;9(1):680–686.

